# The Role of Curcumin Supplementation in Patients with Migraine: A Meta-Analysis of Randomized Controlled Trial

**DOI:** 10.1101/2024.10.29.24316329

**Authors:** Xingke Zhu

## Abstract

**Objectives:** Migraine is a repeated, chronic and neurovascular disease that adversely affects the quality of life and increases the risk of cerebral lesion. Curcumin, the orange-yellow substance of turmeric, may possess anti-headache performance according to several studies. Thus, this study set out to meta-analytically access the anti-headache effect of curcumin supplementation in patients with migraine.

**Methods:** Five databases were searched as of September 30, 2022 to identify all eligible randomized controlled trials. The random-effect Hunter-Schmidt model was used to calculate the effect sizes based on the heterogeneity. The PROSPERO registration number for this meta-analysis is CRD42023409829 (https://www.crd.york.ac.uk/PROSPERO/).

**Results:** Four studies involving 170 patients finally met our inclusion criteria. In the results, curcumin supplementation showed a significant difference in the severity of migraine symptoms compared with placebo (Hedges’s g= -0.75, 95% confidence interval (CI) =-1.44 to -0.07, *P*= 0.03).

**Conclusions:** Curcumin supplementation may relieve the severity of headache symptoms in migraine sufferers.

## Introduction

Migraine is a common disabling primary headache disorder that adversely affects the quality of life^[1]^. It is known as a highly prevalent headache disorder that has a substantial impact on the individual and society^[2]^. Current epidemiological studies, including the GBD study, have shown that migraine is the second leading cause of disability, ranking first among young people, with an estimated global prevalence of 15.2% and 4.9% of all children with disabilities^[3, 4]^. Until now, the understanding of the pathophysiology of migraine is advancing rapidly, and the characterization and diagnosis of its clinical features are constantly improving. These all have led to the view of migraine as a complex and variable disorder of nervous system rather than simply a vascular headache^[5]^. Because of the complexity of the pathophysiological mechanisms of migraine, satisfactory treatment options or highly effective drugs are still limited^[6]^.

Curcumin, a polyphenol, is the main component of the perennial herb Curcuma longa (commonly known as turmeric), which was first isolated in 1815 and has been used for thousands of years^[7, 8]^. Hailed as the “spice for life”, this ingredient from the age-old but ubiquitous Asian plant, turmeric, is now being given a more modern target because of its wide range of physiological activities^[9, 10]^. Toxicity studies have confirmed that curcumin has a good safety profile in long-term ginger use or at high doses (12 g/day), however, its bioavailability is not good enough^[11-13]^. In addition to its antioxidant, anti-inflammatory, anti-bacterial, anti-ageing and anti-cancer activities, curcumin, the “panacea”, has received increasing attention in recent years for its efficacy in neurological diseases^[14]^. A meta-analysis suggested that curcumin might have benefits in improving the symptoms in people with depression^[15]^. And curcumin nanoparticles may treat neurodegenerative diseases, such as Alzheimer’s disease, by enhancing the brain’s self-healing mechanisms^[16]^. Additionally, some clinical trials have preliminarily demonstrated that curcumin supplementation can be used as a treatment for migraine^[17]^.

However, the therapeutic role of curcumin in migraine is still controversial. The evidence in vitro has shown curcumin to be a potential migraine treatment by mitigating H_2_O_2_-induced oxidative stress and cell death in human umbilical vein endothelial cells^[18]^. In an experimental migraine model of rats, the combination of liposome curcumin and sumatriptan showed an improved anti-nociceptive effect^[19]^. Human studies also indicated that curcumin could significantly relieve migraine symptoms^[20, 21]^. But a randomized double-blind, placebo-controlled trial showed that despite a trend for a higher therapeutic effect, no significant relief of headache symptoms was observed in curcumin compared to the placebo group^[22]^. There is still a lack of thorough research on the effects of curcumin on migraine. More evidence is still warranted regarding the optimal dosage, duration and formulation of curcumin for clinical use^[23]^.

Therefore, this research was to evaluate the performance of curcumin or its supplementation in reducing migraine symptoms compared to placebo. A meta-analysis was used to investigate the effect of curcumin on migraine. Three outcomes of migraine symptoms were focused on, including frequency, duration and severity. Besides, subgroup analyses were further used to investigate the differences based on patient’s age, prophylactic administration and dose of curcumin, respectively.

## Methods

We followed the guidelines provided in the Preferred Reporting Items for Systematic Reviews and Meta-analysis, the PRISMA statement, to search the literature and present the results.

### Search strategy

The following databases have been systematically searched for relevant studies from 1984 to March 2023: PubMed, Web of Science, Cochrane Library databases, CNKI and Wanfang Data. A combination of the following keywords was employed to locate relevant studies: (curcuma OR curcuma longa OR curcuminoids OR curcumin OR turmeric) AND (head OR headache OR migraine OR migraines OR cephalalgia OR encephalalgia). All studies were imported into EndNote X9 (Clarivate, Philadelphia, USA) and duplicates were deleted. An initial screening of titles and abstracts for relevance was carried out.

### Inclusion and exclusion criteria

The inclusion criteria were as follows: (1) Only randomized controlled trials were included; (2) Patients who exhibit symptoms of headache; (3) The intervention groups were treated with various formulations of curcumin or curcumin with other drugs. (4) Primary outcomes were the duration, severity and frequency of headache and secondary outcomes were inflammatory indicators.

Also, the exclusion criteria were as follows: (1) Review studies, foundation researches, case studies, guidelines, protocol studies or other unrelated topics; (2) all studies containing the same data were excluded, except for the one with the most representative data; (3) studies without quantitative data referring to the headache symptoms.

### Data extraction and quality assessment

All titles and abstracts of studies that we searched electronically and manually were downloaded. Data were extracted into extraction files by two independent reviewers and discussed and made available by all panel members. Baseline features and target parameters were extracted from selected articles. For every included study, the following characteristics were extracted: (1) Characteristics of studies (the author, year of publication); (2) Characteristics of patients (sample size, gender composition and age); (3) The intervention of experimental and control arms (dosage, formulation, duration); (4) The primary and secondary outcomes described above.

The Jadad scale and Cochrane Collaboration Risk of Bias Tool (CCRBT) was used to evaluate the quality of each observational study^[24]^. The Jadad scoring criteria required the experimenter to generate a proper random sequence (2 points), conduct proper randomization concealment (2 points), use proper blinding (2 points), describe the participants who dropped out (1 point), and a score of 4 or more was good for the quality of the study. The CCRBT was used to evaluate selection bias, performance bias, detection bias, attrition bias, reporting bias and other biases in randomized controlled studies. The ratings “high risk”, “low risk” and “unclear” were used to classify the risk of bias. The Cochrane Collaboration Review Manager software (RevMan) version 5.4.1 (The Nordic Cochrane Centre, The Cochrane Collaboration, Copenhagen) was used for literature bias risk assessment. Two investigators carried out this work separately.

### Assessment of heterogeneity

The heterogeneity was analyzed by I^2^ test. The random-effect model would be used if I^2^ > 50%. The fixed effect model would be used if I^2^ ≤ 50%. When faced with heterogeneity, the subgroup analysis was carried out to explore clinical heterogeneity in terms of interventions and patient types.

### Statistical analysis

Stata16.0 (Stata Corp, College Station, TX, USA) was used for meta-analysis and sensitive analysis. For the primary outcomes (duration, frequency and severity), based on a random effects model, Hedges’s g (an indicator of standardized mean differences suitable for effect size estimation of small sample studies) and 95% confidence intervals (CI) were applied to estimate all comparisons^[25]^. Participants were adults with a current diagnosis of migraine meeting the International Headache Society (HIS) criteria (headache ≥ 15 days per month, lasting more than 3 months or ≥ 1 attack per week)^[26]^. The following continuous variables are analyzed: Duration (average duration of attacks in hours); Frequency (headache frequency per month); Severity (average severity of migraine attacks using the visual analogue scale on a 0–10 numeric scale).

In terms of patient characteristics, there was a lot of evidence that migraine incidence and treatment effectiveness are related to patient’s age and gender^[27-29]^. However, because the included studies lacked individual sex data, grouping based on patient’s sex were not available. Previous studies showed that the efficacy of migraine treatment was related to curcumin dose and whether it was administered prophylactically^[30-32]^. Therefore, we conducted subgroup analyses of the included studies by participants’ age, dosage of curcumin and condition of prophylactic administration respectively.

The types of subgroup meta-analyses were based on age group (< 35 years and ≥ 35 years), prophylactic administration (yes and not), and dose of curcumin (80 mg/day and more than 80 mg/day). The subgroup meta-analyses were carried out separately for outcome indicators of patient symptoms (frequency, duration, severity), following the method of analysis described above.

## Results

### Description of the included studies

The study flow diagram was presented in Fig. 1. Up to September 30, 2022, 507 records were identified by electronic searching in PubMed, Web of Science, Cochrane Library databases, CNKI and Wanfang databases. Seventy-nine duplicate studies and 560 reports not related to population studies were ruled out. No grey literature was found. After reading the titles and abstracts of the remaining 88 studies, 74 studies that met the exclusion criteria were excluded. After reviewing the full text of the remaining studies, 10 studies were excluded due to a lack of data or duplication of data for the primary outcomes. Finally, 4 studies were eligible for this meta-analysis that met all requirements^[21, 33-35]^.

**Figure 1.**
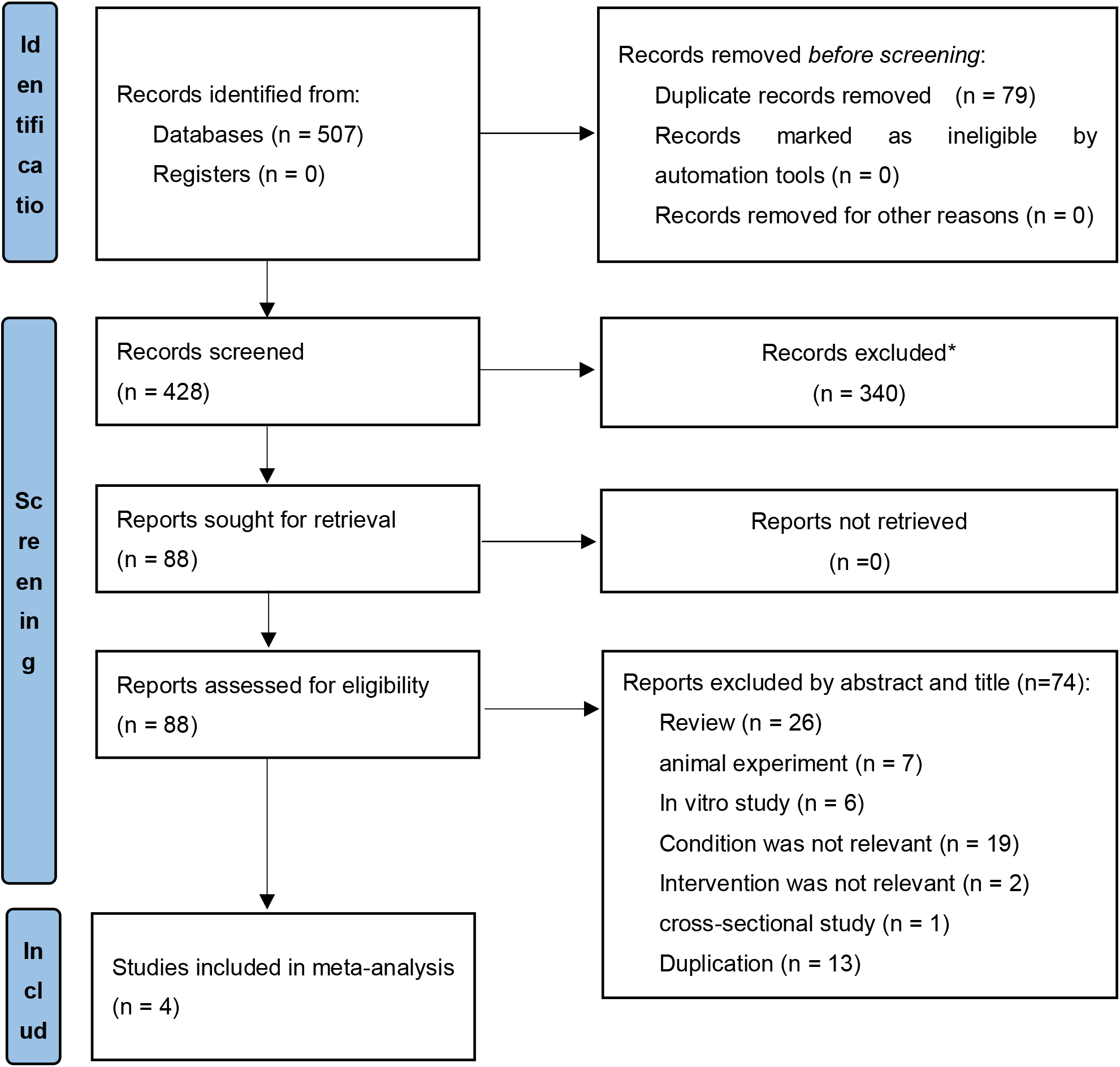
Search strategy diagram. ^*^Reports not related to population studies were excluded by EndnoteX9.

The characteristics of the included trials were summarized in Table 1. Data from the four randomized controlled trials were then pooled. In total, 170 patients were enrolled and completed these studies, including 86 patients in the intervention group and 84 patients in the control group. The included trials were implemented between 2015 and 2021. All of these studies recruited patients diagnosed with migraine according to the previously described standardized tool. Except for one study, preventive medication was used in both intervention and control groups in the remaining studies, while the intervention group also had curcumin supplementation, and the control group had a placebo. All chosen trials quantified migraine symptoms as the primary outcome in different aspects and estimated the effect of intervention using standardized tools. Secondary outcomes were not included in this meta-analysis because of the different data types.

**Table 1.** The characteristics of the included trials.

| Study | Year | Participants<br>(treatment/control) | Age (Mean ± SE) | | Curcumin<br>dosage (mg/day) | Dosage<br>form | Prophylactic<br>medication | Migraine symptoms | | | Other<br>outcomes |
| --- | --- | --- | --- | --- | --- | --- | --- | --- | --- | --- | --- |
|  |  |  | Treatment | Control |  |  |  | Duration | Severity | Frequency |  |
| 1 | 2015 | 19/19 | 37.36±1.95 | 36.57±1.87 | 80 | Nano-cur<br>cumin | Amitriptyline,<br>Propranolol,<br>etc | NS | S | S | VCAM |
| 2 | 2018 | 23/21 | 33.60±1.73 | 31.75±1.89 | 80 | Nano-cur<br>cumin | Topiramate,<br>Amitriptyline,<br>etc | NS | NS | S | None |
| 3 | 2021 | 22/22 | 37.00±1.70 | 37.00±1.50 | 1000 | curcumin<br>pellet | Topiramate | S | S | NS | IL-6,<br>CGRP |
| 4 | 2021 | 22/22 | 39.27±2.15 | 41.00±2.42 | 80 | Nano-cur<br>cumin | None | S | S | S | None |

The intervention time of all studies was 8 weeks.
SE: Standard error. NS: No significant difference compared with the control group. S: Significant difference compared with the control group.

### Studies quality assessments

The studies quality assessments were summarized in Fig. 2. By Jadad criteria, the four included studies all had scores ≥ 4. According to the Cochrane criteria, 75% of the studies elucidated appropriate random sequence generation, 25% performed appropriate allocation concealment, one hundred percent performed appropriate blinding, 50% had complete outcome data, while 50% had selective reporting of risk. These results suggest that the quality of the included studies was generally good, particularly in terms of blinding and randomization compared to other aspects.

**Figure 2.**
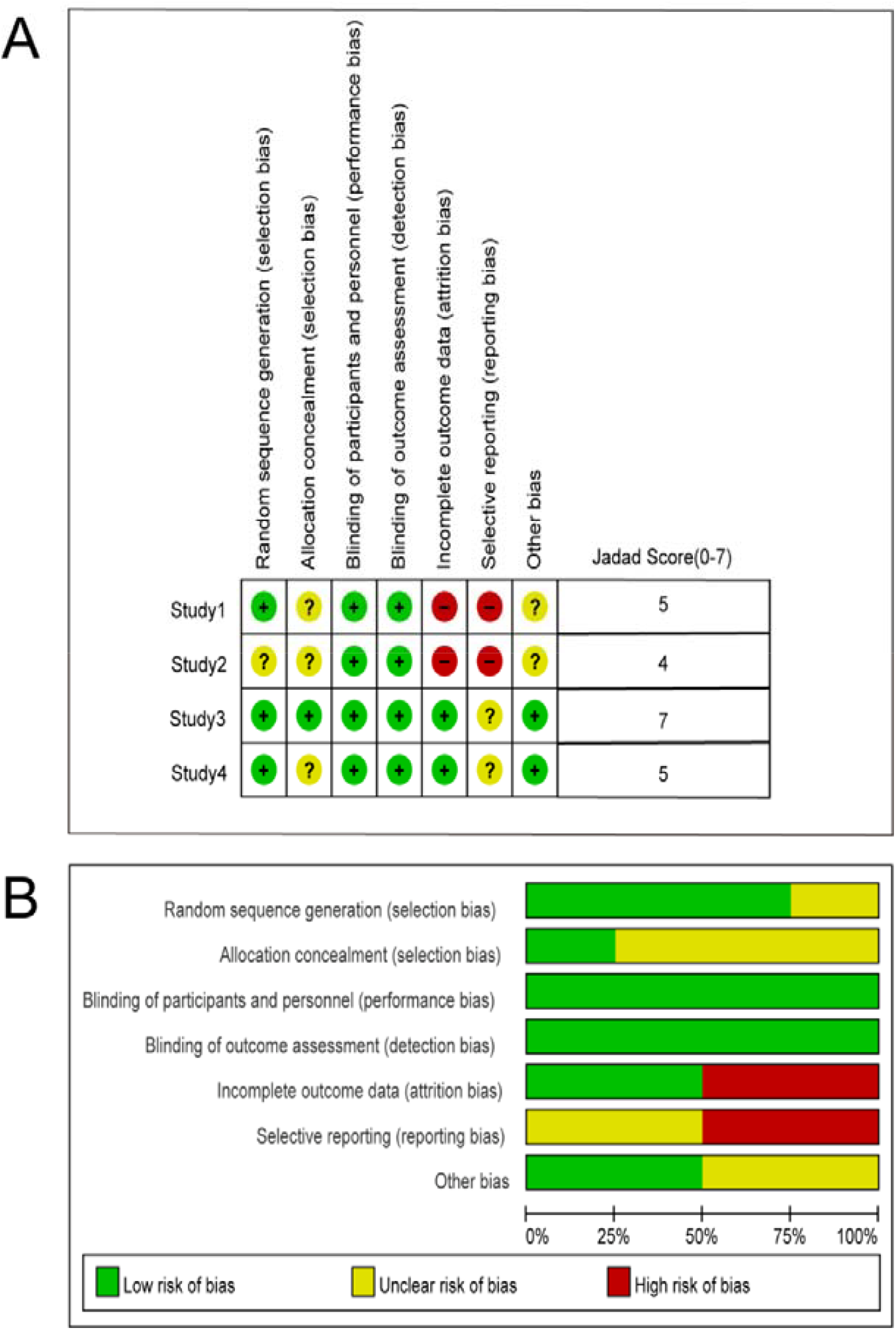
Studies quality assessments. (A) Risk of bias graph for each included study; (B) Risk of bias summary for each included study.

### Curcumin supplementation versus placebo

As shown in Fig. 3, four studies were meta-analyzed separately using frequency, duration and severity as effect sizes. The results indicated that curcumin supplementation showed a significant effect on the severity of migraine reduction (Hedges’s g = -0.75, 95%CI= -1.44 to -0.07, *P*= 0.03). However, significant effects were not observed in the frequency and duration of migraine reduction (for frequency, Hedges’s g= -0.42, 95%CI= -1.15 to 0.32, *P*= 0.27; for duration, Hedges’s g= -0.43, 95%CI= -0.90 to 0.04, *P*= 0.08). According to Cochrane Handbook, the results of duration represented a moderate heterogeneity (I^2^= 58.56%, *P*= 0.02), while the frequency and the severity suggested substantial heterogeneities (I^2^= 82.29%, *P*< 0.01; I^2^=72.08%, *P*< 0.01, respectively).

**Figure 3.**
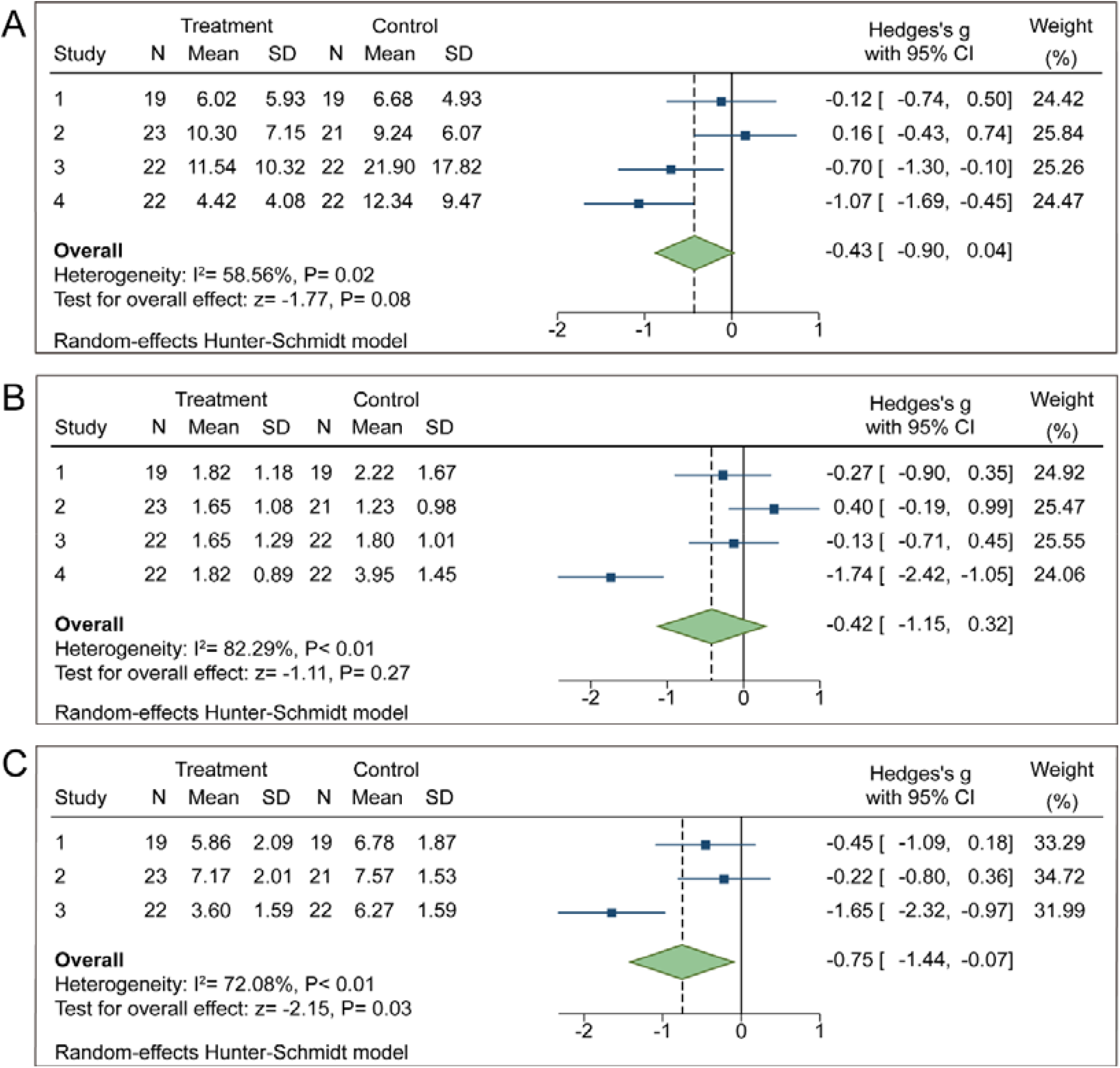
Curcumin supplementation compared to placebo for migraine symptoms. (A) Effect on duration; (B) Effect on frequency; (C) Effect on severity.

Thus, we further conducted the subgroup analyses of these three dimensions (duration, frequency and severity) according to participants’ age, dosage of curcumin, and condition of prophylactic administration respectively.

### Side events

None of the included studies reported any significant adverse effects from the administration of curcumin.

## Discussion

Previous studies lacked quantitative evaluation of the efficacy of curcumin supplementation for migraine. Our study focused on subdividing the onset of migraine symptoms into three aspects (duration, frequency and severity) to investigate the therapeutic effects of curcumin supplementation. Four clinical trials, with a total of 170 patients, were identified to examine the effects of curcumin administration on migraine symptoms. The results suggested that curcumin supplementation was effective in reducing the severity of migraines.

Our findings indicated that curcumin supplementation was effective in reducing the severity of migraine symptoms with significant difference between the control and intervention group. Consistent with previous investigations, curcumin was shown to alleviate the severity of pain. Many population-based trials have proved that curcumin can relieve the painful symptoms of knee osteoarthritis compared to the placebo group^[36, 37]^. There was evidence that curcumin significantly reduced the severity of pain in patients with dysmenorrhea^[38]^. An evaluation of visual analogue scale scores in a meta-analysis suggested that curcumin could relieve the severity of muscle pain ^[39]^. The mechanism of action of curcumin in the treatment of migraine is attributed to its antioxidant and anti-inflammatory properties^[40]^. The relationship among oxidative stress, inflammatory factors (calcitonin gene-related peptide, IL-6, IL-1β, TNF-α, etc.) and migraine has received numerous concerns, although the definitive mechanisms of migraine pathogenesis is still unknown^[41]^. Furthermore, our results indicated that curcumin supplementation did not significantly reduce the frequency and duration of migraine attacks compared to the control group, although results from some of the included studies showed significant differences between the control and intervention groups. It was reasonable that the results might be due to the insufficient sample size and different baseline characteristics of the study objects. Therefore, more studies on curcumin for migraine and larger sample sizes are needed.

The heterogeneity of the subgroup analysis was reduced from moderate to mild, which indicated that age might be a source of heterogeneity in studies of curcumin to reduce migraine duration. Earlier epidemiological studies have also demonstrated that the prevalence of migraine peaks between the ages of thirty and forty and decreases subsequently^[42]^. A previous report of migraine patients revealed a correlation between age and metabolic changes in key regions of the brain^[43]^. Thus, it is reasonable that curcumin supplementation may have better efficacy in the middle-aged population, which may be related to the different prevalence and incidence of migraine in the middle-aged population compared to other age groups.

It is generally accepted that high doses of curcumin are more effective than low doses of curcumin in terms of nerve recovery ^[30, 44]^. Previous studies have shown that the association of low absorption by the small intestine and reductive and conjugative metabolism by the liver diminishes the oral bioavailability of curcumin^[45]^. The natural food-derived compound curcumin is recognized as a safe substance that is non-toxic to humans, especially when administered orally^[46]^. Thus, high-dose administration of curcumin may be more appropriate.

This is the first meta-analysis of the effects of curcumin supplementation on migraine sufferers, which has explored the efficacy of curcumin through three different dimensions of migraine symptoms. And this meta-analysis provides reasonable suggestions on the appropriate population and dosage of curcumin for migraine. Through the quality assessment, we judged that the included randomized controlled trials were generally of good quality. All included trials described the placebo-controlled design, selection process, patient allocation, and reasons for exiting and met the requirements in terms of blind design. While examining our findings, however, some limitations should be considered. Firstly, due to limited data and requirements for data types, the number of included studies and sample size were small. However, because the studies we included were all randomized double-blind, placebo-controlled trials, the results are plausible. Then, the inflammatory factor indicators were not included in this meta-analysis due to the inconsistent data types. Another limitation have concerned the literature search. Studies published in languages other than English and Chinese or unpublished clinical studies were not included, because the databases these two languages were considered to contain a comprehensive range of clinical studies.

## Conclusions

The findings of this study suggested that curcumin supplementation was effective in reducing the severity of migraines. Clinical studies with larger sample sizes are needed to further validate the appropriate population characteristics and dosage of curcumin for the treatment of migraine.

## Data Availability

All data produced in the present study are available upon reasonable request to the authors

https://www.crd.york.ac.uk/PROSPERO/

## References

[1] Headache Classification Committee of the International Headache Society (IHS).The International Classification of Headache Disorders, 3rd edition (beta version). Cephalalgia 2013;33(9):629–808.

[2] Bigal ME, Lipton RB, Stewart WF. The epidemiology and impact of migraine. Curr Neurol Neurosci Rep 2004;4(2):98–104.

[3] Steiner TJ, Stovner LJ. Global epidemiology of migraine and its implications for public health and health policy. Nat Rev Neurol 2023;19(2):109–17.

[4] GBD 2019 Diseases and Injuries Collaborators. Global burden of 369 diseases and injuries in 204 countries and territories, 1990-2019: a systematic analysis for the Global Burden of Disease Study 2019. Lancet 2020;396(10258):1204–22.

[5] Charles A. The pathophysiology of migraine: implications for clinical management. Lancet Neurol 2018;17(2):174–82.

[6] Diener HC, Charles A, Goadsby PJ, et al. New therapeutic approaches for the prevention and treatment of migraine. Lancet Neurol 2015;14(10):1010–22.

[7] Goel A, Kunnumakkara AB, Aggarwal BB. Curcumin as “Curecumin”: from kitchen to clinic. Biochem Pharmacol 2008;75(4):787–809.

[8] Kotha RR, Luthria DL. Curcumin: Biological, Pharmaceutical, Nutraceutical, and Analytical Aspects. Molecules 2019;24(16).

[9] Balachandran K, Stebbing J. Turmeric: a spice for life? Lancet Oncol 2016;17(12):1639.

[10] Aggarwal BB, Sung B. Pharmacological basis for the role of curcumin in chronic diseases: an age-old spice with modern targets. Trends Pharmacol Sci 2009;30(2):85–94.

[11] Pivari F, Mingione A, Piazzini G, et al. Curcumin Supplementation (Meriva(®)) Modulates Inflammation, Lipid Peroxidation and Gut Microbiota Composition in Chronic Kidney Disease. Nutrients 2022 ;14(1).

[12] Patel SS, Acharya A, Ray RS, et al. Cellular and molecular mechanisms of curcumin in prevention and treatment of disease. Crit Rev Food Sci Nutr 2020;60(6):887–939.

[13] Lao CD, Ruffin MTt, Normolle D, et al. Dose escalation of a curcuminoid formulation. BMC Complement Altern Med 2006 ;6:10.

[14] Um MY, Yoon M, Kim M, et al. Curcuminoids, a major turmeric component, have a sleep-enhancing effect by targeting the histamine H1 receptor. Food Funct 2022;13(24):12697–706.

[15] Fusar-Poli L, Vozza L, Gabbiadini A, et al. Curcumin for depression: a meta-analysis. Crit Rev Food Sci Nutr 2020;60(15):2643–53.

[16] Tiwari SK, Agarwal S, Seth B, et al. Curcumin-loaded nanoparticles potently induce adult neurogenesis and reverse cognitive deficits in Alzheimer’s disease model via canonical Wnt/β-catenin pathway. ACS Nano 2014;8(1):76–103.

[17] Lopresti AL, Smith SJ, Drummond PD. Herbal treatments for migraine: A systematic review of randomised-controlled studies. Phytother Res 2020;34(10):2493–517.

[18] Ouyang J, Li R, Shi H, et al. Curcumin Protects Human Umbilical Vein Endothelial Cells against H_2_O_2_-Induced Cell Injury. Pain Res Manag 2019;2019:3173149.

[19] Bulboacă AE, Bolboacă SD, Stănescu IC, et al. The effect of intravenous administration of liposomal curcumin in addition to sumatriptan treatment in an experimental migraine model in rats. Int J Nanomedicine 2018;13:3093–103.

[20] Martin BR. Multimodal Care for Headaches, Lumbopelvic Pain, and Dysmenorrhea in a Woman With Endometriosis: A Case Report. J Chiropr Med 2021;20(3):148–57.

[21] Sedighiyan M, Abdolahi M, Jafari E, et al. The effects of nano-curcumin supplementation on adipokines levels in obese and overweight patients with migraine: a double blind clinical trial study. BMC Res Notes 2022;15(1):189.

[22] Askari G, Sahebkar A, Soleimani D, et al. The efficacy of curcumin-piperine co-supplementation on clinical symptoms, duration, severity, and inflammatory factors in COVID-19 outpatients: a randomized double-blind, placebo-controlled trial. Trials 2022;23(1):472.

[23] Ferguson JJA, Abbott KA, Garg ML. Anti-inflammatory effects of oral supplementation with curcumin: a systematic review and meta-analysis of randomized controlled trials. Nutr Rev 2021;79(9):1043–66.

[24] Lin Y, Chen Y, Liu R, et al. Effect of exercise on rehabilitation of breast cancer surgery patients: A systematic review and meta-analysis of randomized controlled trials. Nurs Open 2023;10(4):2030–2043.

[25] Therese D. Pigott. Methods of meta-analysis: Correcting error and bias in research findings - ScienceDirect. Evaluation and Program Planning 2006;29(3):236–237.

[26] Ward TN. Migraine diagnosis and pathophysiology. Continuum (Minneap Minn) 2012;18(4):753–63.

[27] Lebedeva ER. Sex and age differences in migraine treatment and management strategies. Int Rev Neurobiol 2022;164:309–47.

[28] Vetvik KG, MacGregor EA. Sex differences in the epidemiology, clinical features, and pathophysiology of migraine. Lancet Neurol 2017;16(1):76–87.

[29] Ahmad SR, Rosendale N. Sex and Gender Considerations in Episodic Migraine. Curr Pain Headache Rep 2022;26(7):505–16.

[30] Luo Y, Huang Q, Chen D. The Pharmacodynamic Study of Curcumin on Chronic Atrophic Gastritis in Rats. Journal of Jiangxi University of Chinese Medicine 2012;24(04):58–60.

[31] Brandes JL, Saper JR, Diamond M, et al. Topiramate for migraine prevention: a randomized controlled trial. JAMA 2004;291(8):965–73.

[32] Diener HC, Tfelt-Hansen P, Dahlöf C, et al. Topiramate in migraine prophylaxis--results from a placebo-controlled trial with propranolol as an active control. J Neurol 2004;251(8):943–50.

[33] Abdolahi M, Karimi E, Sarraf P, et al. The omega-3 and Nano-curcumin effects on vascular cell adhesion molecule (VCAM) in episodic migraine patients: a randomized clinical trial. BMC Res Notes 2021;14(1):283.

[34] Parohan M, Sarraf P, Javanbakht MH, et al. The synergistic effects of nano-curcumin and coenzyme Q10 supplementation in migraine prophylaxis: a randomized, placebo-controlled, double-blind trial. Nutr Neurosci 2021;24(4):317–26.

[35] Rezaie S, Askari G, Khorvash F, et al. Effects of Curcumin Supplementation on Clinical Features and Inflammation, in Migraine Patients: A Double-Blind Controlled, Placebo Randomized Clinical Trial. Int J Prev Med 2021;12:161.

[36] Lopresti AL, Smith SJ, Jackson-Michel S, et al. An Investigation into the Effects of a Curcumin Extract (Curcugen^®^) on Osteoarthritis Pain of the Knee: A Randomised, Double-Blind, Placebo-Controlled Study. Nutrients 2021;14(1).

[37] Jamali N, Adib-Hajbaghery M, Soleimani A. The effect of curcumin ointment on knee pain in older adults with osteoarthritis: a randomized placebo trial. BMC Complement Med Ther 2020;20(1):305.

[38] Bahrami A, Zarban A, Rezapour H, et al. Effects of curcumin on menstrual pattern, premenstrual syndrome, and dysmenorrhea: A triple-blind, placebo-controlled clinical trial. Phytother Res 2021;35(12):6954–62.

[39] Beba M, Mohammadi H, Clark CCT, et al. The effect of curcumin supplementation on delayed-onset muscle soreness, inflammation, muscle strength, and joint flexibility: A systematic review and dose-response meta-analysis of randomized controlled trials. Phytother Res 2022;36(7):2767–78.

[40] Goschorska M, Gutowska I, Baranowska-Bosiacka I, et al. The Use of Antioxidants in the Treatment of Migraine. Antioxidants (Basel) 2020;9(2).

[41] Ramachandran R. Neurogenic inflammation and its role in migraine. Semin Immunopathol 2018;40(3):301–14.

[42] Manzoni GC, Stovner LJ. Epidemiology of headache. Handb Clin Neurol 2010;97:3–22.

[43] Lisicki M, D’Ostilio K, Coppola G, et al. Age related metabolic modifications in the migraine brain. Cephalalgia 2019;39(8):978–87.

[44] Yao M, Yang L, Wang J, et al. Neurological recovery and antioxidant effects of curcumin for spinal cord injury in the rat: a network meta-analysis and systematic review. J Neurotrauma 2015;32(6):381–91.

[45] Dei Cas M, Ghidoni R. Dietary Curcumin: Correlation between Bioavailability and Health Potential. Nutrients 2019;11(9).

[46] Soleimani V, Sahebkar A, Hosseinzadeh H. Turmeric (Curcuma longa) and its major constituent (curcumin) as nontoxic and safe substances: Review. Phytother Res 2018;32(6):985–95.

